# Discovery of runs-of-homozygosity diplotype clusters and their associations with diseases in UK Biobank

**DOI:** 10.1101/2020.10.26.20220004

**Authors:** Ardalan Naseri, Degui Zhi, Shaojie Zhang

**Affiliations:** Department of Computer Science, University of Central Florida, Orlando, Florida 32816, USA; Center for Precision Health, School of Biomedical Informatics, University of Texas Health Science Center at Houston, Houston, Texas 77030, USA

**Author notes:** School of Biomedical Informatics, University of Texas Health Science Center at Houston, Houston, Texas 77030, USA.

## Abstract

Runs of homozygosity (ROH) segments, contiguous homozygous regions in a genome were traditionally linked to families and inbred populations. However, a growing literature suggests that ROHs are ubiquitous in outbred populations. Still, most existing genetic studies of ROH in populations are limited to aggregated ROH content across the genome, which does not offer the resolution for mapping causal loci. This limitation is mainly due to a lack of methods for efficient identification of shared ROH diplotypes. Here, we present a new method, ROH-DICE, to find large ROH diplotype clusters, sufficiently long ROHs shared by a sufficient number of individuals, in large cohorts. ROH-DICE identified over 1 million ROH diplotypes that span over 100 SNPs and shared by more than 100 UK Biobank participants. Moreover, we found significant associations of clustered ROH diplotypes across the genome with various self-reported diseases, with the strongest associations found between the extended HLA region and autoimmune disorders. We found an association between a diplotype covering the HFE gene and haemochromatosis, even though the well-known causal SNP was not directly genotyped nor imputed. Using genome-wide scan, we identified a putative association between carriers of an ROH diplotype in chromosome 4 and an increase of mortality among COVID-19 patients. In summary, our ROH-DICE method, by calling out large ROH diplotypes in a large outbred population, enables further population genetics into the demographic history of large populations. More importantly, our method enables a new genome-wide mapping approach for finding disease-causing loci with multi-marker recessive effects at population scale.

## Introduction

Runs of homozygosity (ROH) regions are regions of diploid chromosomes where identical-by-descent (IBD) haplotypes are inherited from each parent^1^. Traditionally, ROH were thought to be relevant only to inbred populations, and ROH may link to consanguinity and population isolation^2^. However, a growing number of studies of large cohorts and biobanks have found that ROH may be ubiquitously present^3,4^. Still, our understanding of the genetic impacts of ROH are limited.

Most existing studies used individuals’ global ROH content (the sum of lengths or the count of ROHs) as a surrogate for the degree of inbreeding and associated it with phenotypes. It has long been known that inbreeding is harmful to the health of offspring^5^, and several studies have suggested that the global ROH content is associated with higher risks of recessive disorders^6–8^. ROHs can also be related to complex traits such as height^9^. With the growing trend of multi-cohort collaboration through meta-analysis, the effect of global ROH content has been studied over very large sample sizes^3,4^. A recent study^10^ revealed that people with extremely long ROH can be found even in outbred populations.

However, collapsing the individual’s rich ROH content into a single number summarizing their global content is a drastic oversimplification. In doing so, the opportunities for mapping causal loci of phenotypes are lost. Ideally, one might wish to identify chromosomal regions with a certain ROH diplotype^11^ (pairs of identical haplotypes), and associate the ROH diplotype with the phenotypes of interest. Indeed, homozygosity mapping in pedigree or inbred population has achieved success in identifying recessive loci^7,12–16^. However, for general outbred populations, the total number of possible ROH diplotypes at a locus are too enormous to be enumerated efficiently, and ROH mapping of outbred population has remained only a theoretical possibility.

Here, we proposed an approach that bypasses this impossibly large search space of diplotypes. Instead of enumerating all ROH diplotype, we focused on those that are sufficiently long and frequent. Such ROH diplotypes are of interest because they are at the extreme of distribution: the chance of ROH is determined by the chance of a pair of mates having IBD, and such chance and also the length of IBD segments will decay quickly in outbred populations, as supported by population genetics theory^17,18^ and real world data^19,20^. However, little is known about such ROH diplotypes because no existing methods can efficiently find them. We present an efficient PBWT-based^21^ method to find clusters of identical matches. We apply our method to find clusters of ROH diplotypes in UK-Biobank data. Each cluster of ROH diplotypes is defined as a set of 100 consecutive homozygous sites that is shared among over 100 individuals. We investigate the association between the detected ROH diplotype clusters and self-reported non-cancerous diseases and present the results for the disease having the strongest associations with the detected ROH diplotype clusters.

## Results

### Methods overview

An ROH diplotype is a pair of homozygous haplotypes of an individual. A frequent ROH diplotype is one shared by a number of individuals at the same location and with the same consensus sequence. Roughly, the frequency of an ROH diplotype would decrease exponentially with its length. Although long and frequent ROH diplotypes are not very common, it is difficult to enumerate ROH diplotypes above a certain length and above a certain frequency. As a compromise, ROH regions are traditionally aggregated into single numbers and their association with phenotypes is investigated. As a result, the loci specific association signals of the ROHs or the allele specific signals are likely to be lost (see **Figure 1**).

**Figure 1:**
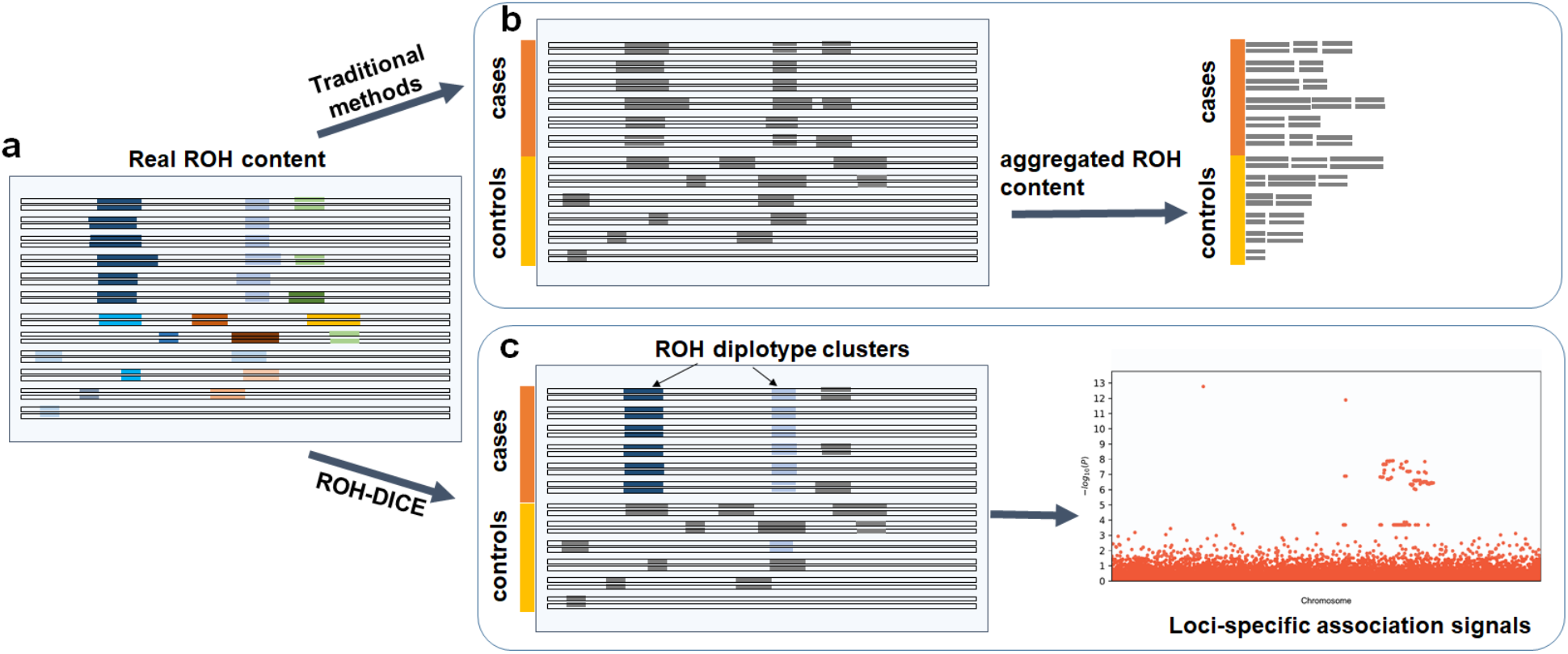
ROH-DICE enables discovery of loci-specific association signals of ROH diplotypes. The actual ROH contents (a) including the locations and sequence identities of ROH (indicated by different colors) were lost in traditional ROH analysis pipelines which (b) aggregate the ROH contents per individual and lose the chances for identifying associating loci. ROH-DICE reveals (c) ROH diplotype clusters that are long and wide enough, thus enabling mapping loci associated with phenotypes.

To solve this problem, we first processed the biallelic genotype panel (with three possible values 0, 1, 2 at each position) by randomly assigning any heterozygous sites into homozygous sites with the reference or the alternative allele. The reasons for such processing are two-folded. One, through this conversion, the true ROH diplotype clusters, mostly consisting of homozygous sites, are relatively intact and will still have a high probability of maintaining a good portion of their haplotype. However, some post hoc processing may be needed to merge the ROH diplotype clustered with minor deviations of their consensus sequences. Notably, this conversion should introduce very few false positives as when the length cutoff and the width cutoff is large, there is little chance a non-ROH diplotype cluster will emerge. Two, this effectively converts the panel into a haplotype panel (with two possible values 0 and 2 at each position), where efficient algorithms for identifying haplotype matching blocks are available. An extra benefit is, by doing this conversion, no phasing of haplotypes is needed.

Haplotype matching blocks can be identified by leveraging the efficient sorting of haplotypes in the Positional Burrows Wheeler transform (PBWT) data structure. For a haplotype panel, PBWT sorts haplotype sequences at each variant site according to their reverse suffixes, and thus a set of haplotypes sharing a same sequence before a variant site will be adjacent in the sorting and form a “matching block”. We use auxiliary PBWT data structures keeping track of the length (the number of variant sites) and the width (the number of haplotype sequences) of the matching block and trigger the output report by watching the data structures. **Figure 2** summarizes the overall ROH-DICE method. More details about the algorithms for finding blocks of matches and searching for ROH diplotypes are presented in the Methods section.

**Figure 2:**
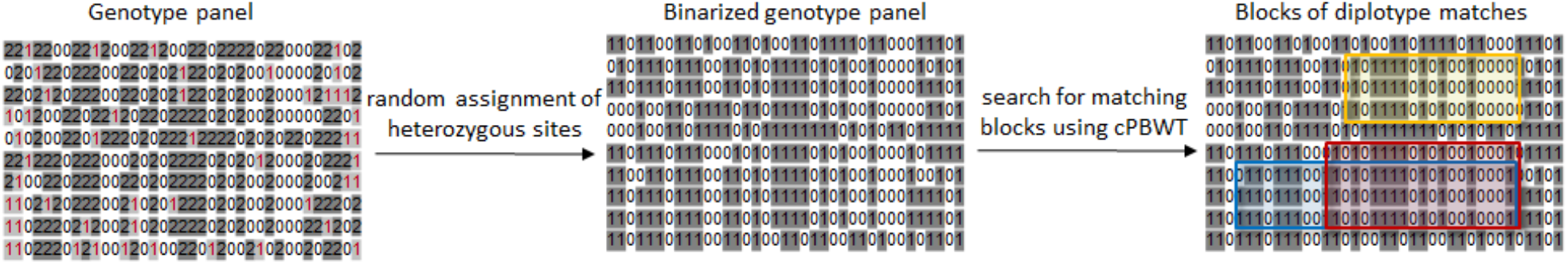
A simple schematic of searching for ROH diplotype clusters in a genotype panel. The input is a genotype panel where each line represents an individual. The heterozygous sites are depicted in violet in the genotype panel. Input genotype data are converted into a binarized genotype panel where homozygous sire are preserved. The matching blocks (clusters) are searched using cPBWT. A matching block defined by a minimum number of sites, individuals and also an objective function. The objective can be either maximizing the number of individuals or maximizing the number of sites. The clusters of matches are highlighted in different colors. Red represents a cluster with the maximized number of individuals and blue represents a cluster with the maximized number of sites.

### ROH diplotypes in UK Biobank

Here, we searched for the clusters of ROH regions in the UK-Biobank data^22^. All autosomal chromosomes of all UK participants (487,409) were searched for ROH regions that are shared among at least 100 individuals comprising at least 100 consecutive sites. 56,972 people with self-reported non-British ethnicity in UK-Biobank were filtered out. On average ∼18% of sites are heterozygous, and thus for a pair of 100 sites genotype sequences, there is a very small probability that they will be mapped to the same compressed haplotype. Thus, the rate of false positive should be low. To increase statistical power for downstream association tasks, the width-maximal blocks were reported. This was achieved by running the ROH-DICE program, which took 15 CPU days and 158 MB peak memory. After running the ROH-DICE program, further post-processing steps were conducted. Each individual with more than 1% heterozygous sites within the block was removed from the cluster. Any two clusters with the same consensus and the exact starting and ending positions were merged.

A total of 1,880,826 ROH clusters (shared among at least 100 individuals and extending at least 100 consecutive sites) were identified in all 22 autosomal chromosomes. The average length of these ROHs is 553,095 bp (∼0.55 cM). The distribution of ROH clusters is very uneven (**Figure 3**). Interestingly, the number of ROH clusters in chromosome 6 is the highest. This is mainly due to the excessive number of ROH clusters in the MHC region (65,458). **Figure 4** illustrates the genome-wide coverage of the ROH clusters, with visible peaks at chromosomes 2, 6, and 8. A peak region in chromosome 2 (chr2:135755899-136827560) has been reported to harbor a high selection signal^23^. This region contains the LCT gene which includes a variant selected for lactose tolerance in European population^24^. The most prominent peak in chromosome 6 is located in the MHC region (chr6:28,477,797-33,448,354), whose details are shown in **Figure 4**. Surprisingly, some clusters comprise more than a hundred thousand individuals sharing the same ROH consensus. The high rate of ROH clusters in the MHC region may be due to high marker density and low recombination rates^25,26^. The ROHs are abundant in regions with low recombination rates and also their distribution is expected to be population-specific and the hotspots and coldspots may vary in different populations^27^. The ROH coldspot in European population is reported to be located in chromosome 18^27^ which is consistent with our ROH diplotypes results using British individuals in the UK Biobank data. A hotspot was reported in chromosome 15 for Europeans^27^ which overlaps with a peak for British people in chromosome 15 (72100881-72681976).

**Figure 3:**
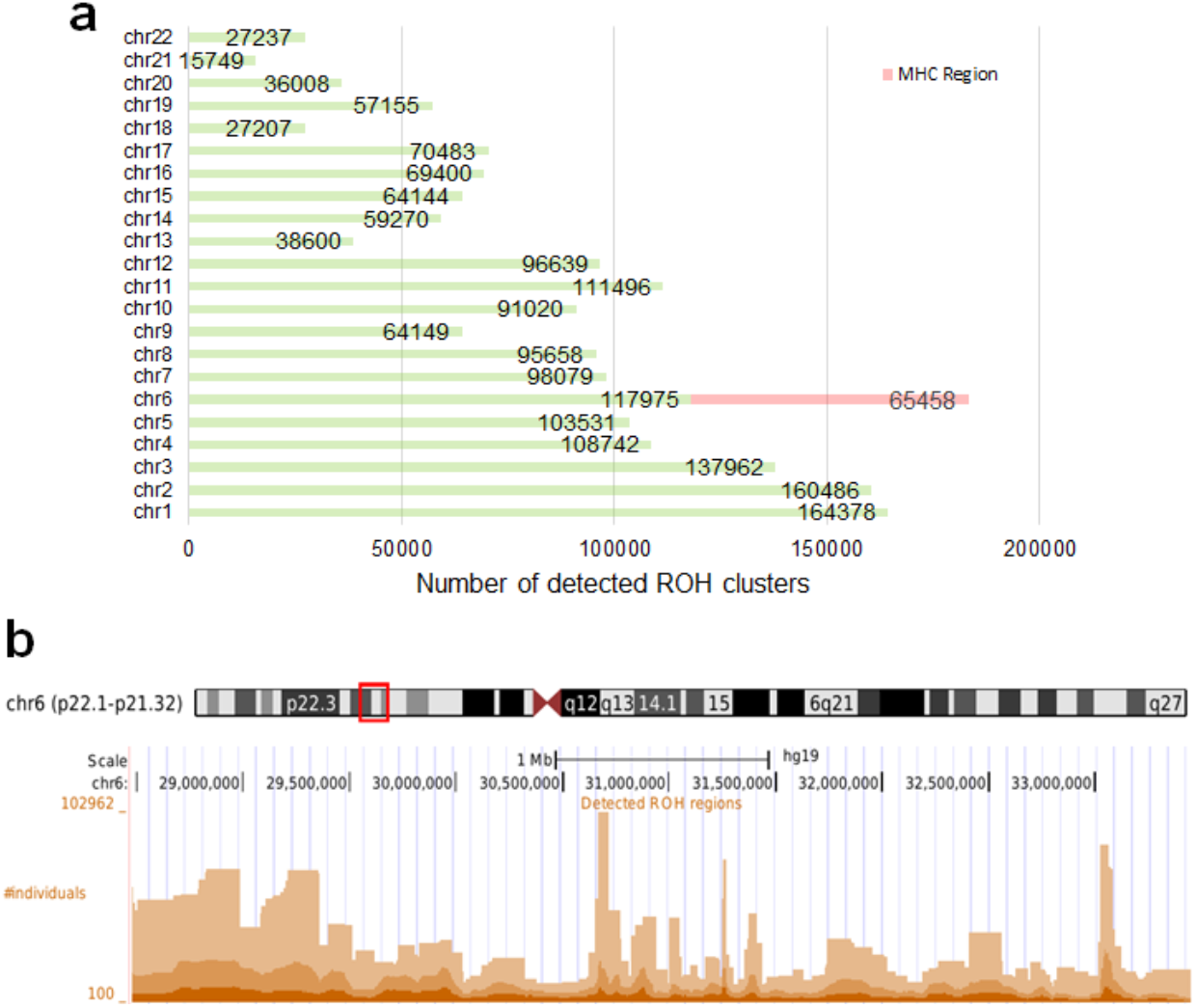
Total number of detected ROH diplotype clusters in each autosomal chromosome (a) and the detected ROH clusters in the MHC region (chr6:28,477,797-33,448,354) in hg19 (b). Some regions may contain multiple overlapping clusters comprising different sets of individuals. The minimum length of the ROH regions was set to 100 sites and the minimum number of individuals to 100.

**Figure 4:**
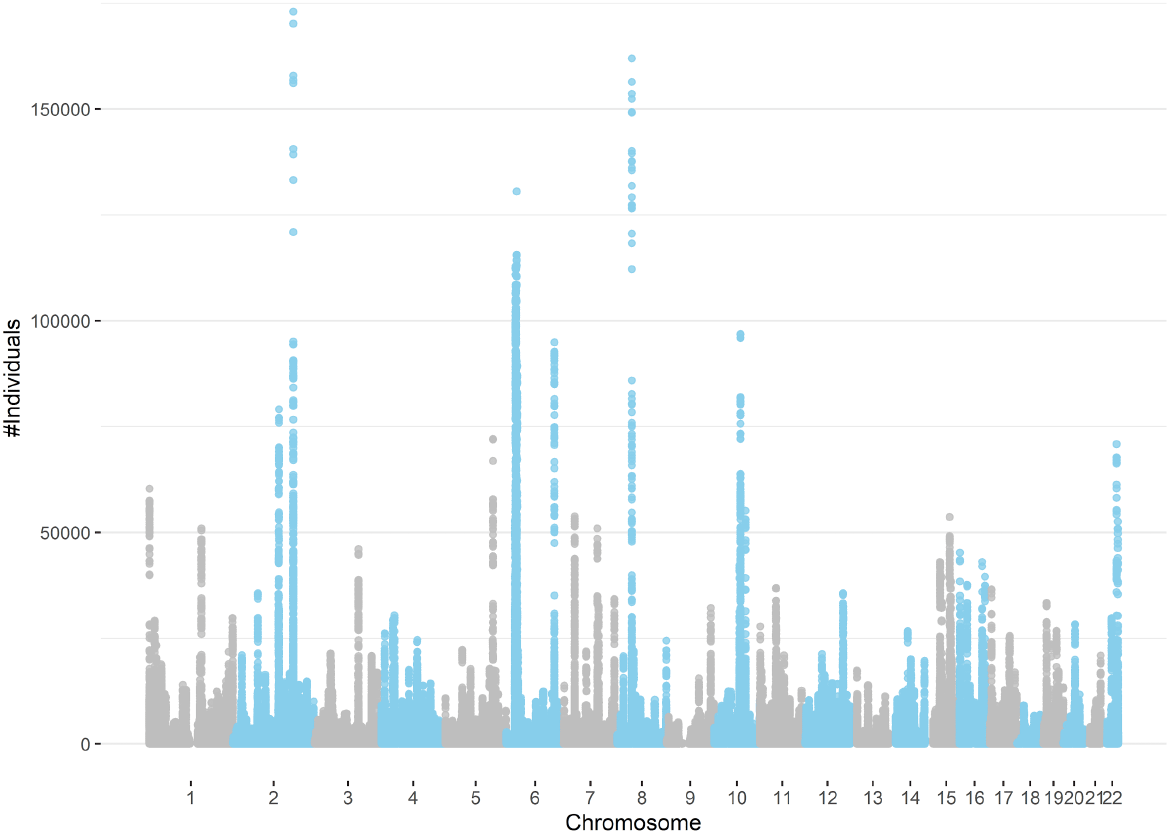
Detected ROH diplotype clusters with at least 100 individuals sharing the same consensus with a minimum number of 100 SNPs. Chromosome 18 has the lowest peak for individuals sharing an ROH diplotype. Chromosomes 2, 6, and 8 contain diplotypes shared with more than 100,000 individuals.

### ROH clusters and disease association

We conducted a phenotypic association analysis of the found ROH diplotype clusters with 445 self-reported non-cancerous diseases, as they are conveniently available in the UK Biobank. We first conducted a quick Chi-squared test associating each of the 1,880,826 ROH diplotype cluster membership against each of the 445 phenotypes (See **Methods**). The p-values for the 100 regions with the lowest p-values were re-computed using age, sex, genetics principal components and genotype measurement batch fields by PHESANT^28^ (Details see **Methods**). This identified 61 associations passing Bonferroni-corrected p-value threshold of 10^−12^. Table 1 shows the p-values for disease associated with the HLA region (chr6) computed by PHESANT. P-values for some diseases are very low in both chi-squared test and *regression* analysis using PHESANT. It also includes the SNP with the lowest p-value in each cluster that is associated with the corresponding disease. The SNP with the lowest p-value in each cluster was extracted from Neale’s lab results^a^. Most of the clusters with low p-value contain at least one SNPS with very low p-value that is associated with the corresponding disease. Top 100 diplotypes with the lowest p-values using chi-squared tests and PHESANT are included in **Supplementary Table 1**.

**Table 1:** Clusters of the ROH diplotypes with the lowest p-values in HLA region for self-reported diseases using the British population in UK-Biobank. The p-values were calculated using PHESANT. For each disease only the region with the lowest p-value has been included.

| disease (binary trait) | position (on chr6) | p-value | carrier frequency | odds ratio | genetic length (cM) | GWAS p-value* | GWAS lead SNP* |
| --- | --- | --- | --- | --- | --- | --- | --- |
| **ankylosing spondylitis** | 31431031-31464050 | $4.62 \times 10^{-34}$ | 0.29% | 8.66 | 0.071198 | 0 | rs113340460 |
| **haemochromatosis** | 25969631-26108168 | $8.02 \times 10^{-120}$ | 0.09% | 24.51 | 0.011597 | - | - |
| **malabsorption/ coeliac disease** | 32564985-32629755 | $3.41 \times 10^{-259}$ | 4.12% | 1.64 | 0.005408 | 0 | rs9271352 |
| **multiple sclerosis** | 32410215-32554129 | $4.36 \times 10^{-45}$ | 0.37% | 3.79 | 0.012736 | $1.05 \times 10^{-107}$ | rs9268925 |
| **polymyalgia rheumatica** | 31710968-31794592 | $7.31 \times 10^{-09}$ | 0.23% | 5.90 | 0.006808 | $1.59 \times 10^{-08}$ | rs1150748 |
| **prostate problem (not cancer)** | 34607958-35163974 | $2.84 \times 10^{-08}$ | 0.18% | 6.94 | 0.034889 | $9.81 \times 10^{-04}$ | rs76117834 |
| **psoriasis** | 31254263-31263216 | $1.20 \times 10^{-122}$ | 1.21% | 2.73 | $3.07 \times 10^{-05}$ | 0 | rs13214872 |
| **psoriatic arthropathy** | 33072522-33115762 | $8.54 \times 10^{-12}$ | 0.20% | 3.97 | 0.008708 | $4.76 \times 10^{-10}$ | rs17221401 |
| **rheumatoid arthritis** | 32412539-32573760 | $8.15 \times 10^{-122}$ | 0.23% | 2.34 | 0.01293 | $6.96 \times 10^{-124}$ | rs188575117 |

\*p-values are for the reported SNP from:
http://www.nealelab.is/blog/2017/9/15/heritability-of-2000-traits-and-disorders-in-the-uk-biobank

Not surprisingly, the most prominent associations we found are ROH diplotype in the HLA region with autoimmune diseases. We found that *malabsorption/coeliac disease, psoriasis, rheumatoid arthrit*is, and *multiple sclerosis* have the strongest association with loci in the HLA region. These results are largely consistent with known literature^29–34^. One of the most significant associations we identified is the association between the ROH diplotype at chr6:25988167-26122453 and haemochromatosis (p-value = 9.16 × 10^−120^). The frequency of the ROH diplotype is only 0.02% and the odds ratio of having the disease for the carrier is 102.21. Interestingly, several other ROH diplotypes at this locus also have strong association with haemochromatosis (**Table 1**). This locus is in the extended HLA region and has a low recombination rate. *Haemochromatosis* is an inherited disorder in which iron levels in the body slowly build up over several years. The gene HFE (chr6:26,087,509-26,095,469) is a well-known recessive locus for this disease^35^. The C282Y polymorphism (rs1800562, chr6:26,092,913) in HFE is the most-penetrant but other polymorphisms with lesser penetrance are also known. Interestingly, the minor allele frequency for the SNP rs1800562 is of 6% allele frequency in the European population but it is not genotyped (and is also not available in the imputed panel) in the UK Biobank data. As a result, this association signal has been completely missing in the Neale Lab results. In another study the SNP has been imputed and a specific association study for the recessive effect between the homozygous alleles of rs1800562 and hemochromatosis has been reported^36^. Basically, our approach found this recessive association signal without direct genotyping of any SNP with high LD to the causal SNP, demonstrating the power of our approach beyond regular additive effect GWAS.

We also found some loci outside of the HLA region that are presumably associated with non-cancerous diseases (**Table 2**). Most prominent one is an ROH diplotype at chr1:151515188-151902494 with eczema/dermatitis. This signal overlaps with the GWAS finding of rs4845604 at chr1:151829204^37^.

**Table 2:**
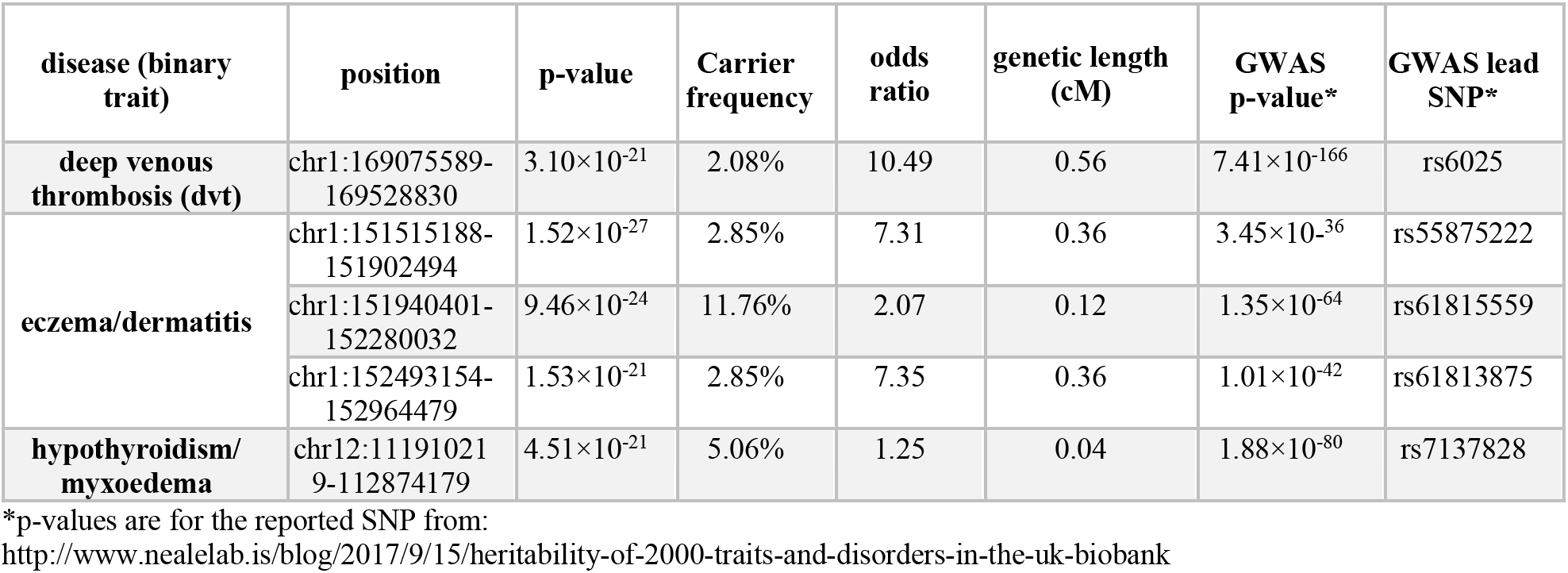
Clusters of the ROHs with the lowest p-values outside of HLA region for self-reported diseases using the British population in UK-Biobank. The p-values were calculated using PHESANT.

### ROH clusters and COVID-19 association

We computed the p-value using the Chi-square test for association between mortality of COVID-19 and the detected ROH regions. We considered only the clusters that had at least 10 cases (tested positive and passed away in 2020). **Figure 5** shows the Manhattan plot for ROH regions and mortality of COVID-19. The most significant ROH region is located in chr4:106318456-106483898 with p-value 1.63×10^−10^. 4,389 individuals share the diplotypes and 76 of them have been tested positive for COVID-19. 11 persons who carried the same ROH consensus and had been tested positive, died in 2020. The region includes the PPA2 gene. The gene product is an inorganic pyrophosphatase located in the mitochondrion^38^. Missense mutations in this gene is reported to cause sudden unexpected cardiac arrest in infancy^39^.

**Figure 5:**
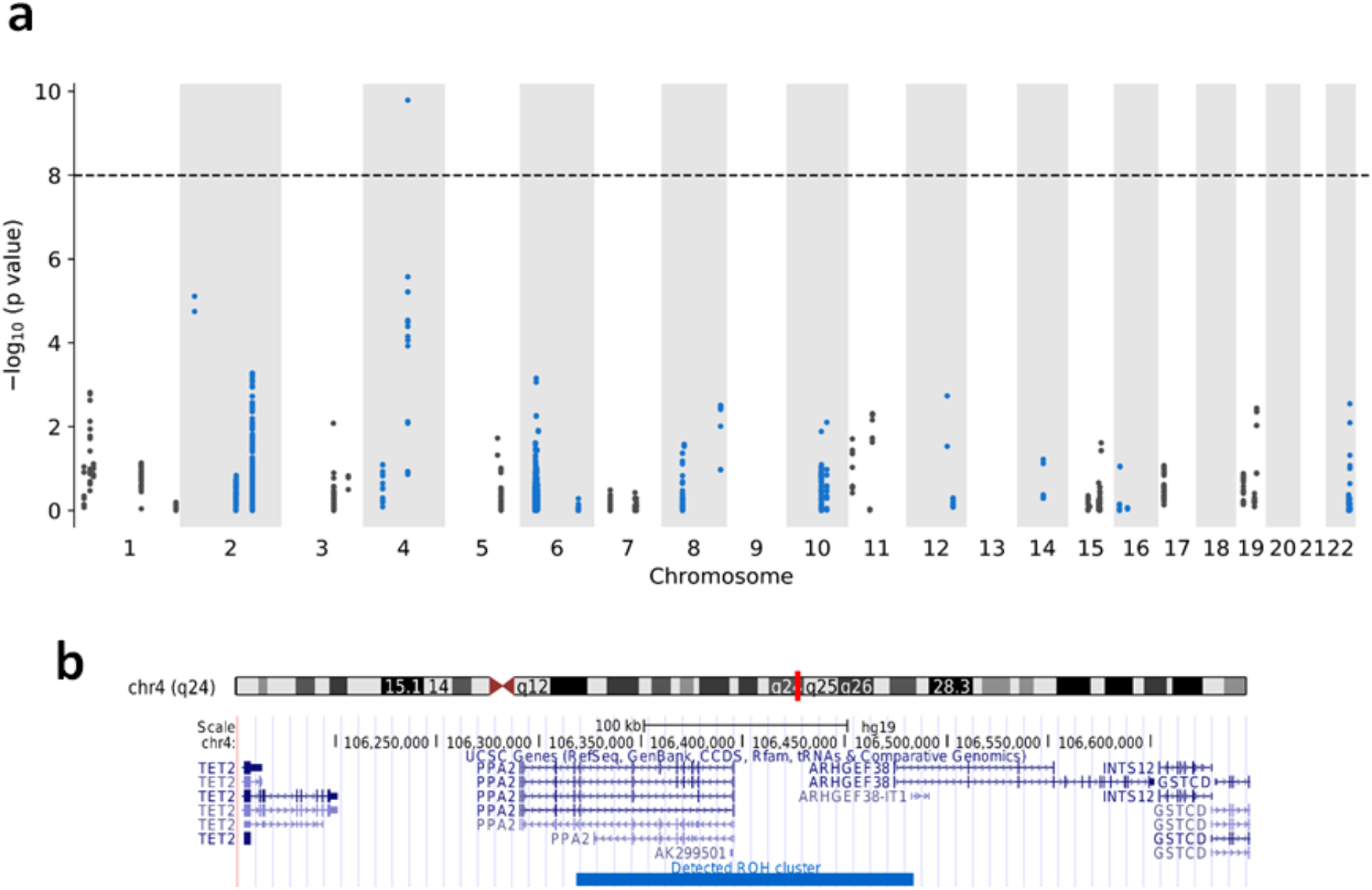
ROH associations between ROH diplotypes and mortality of COVID-19. (a) Manhattan plot of ROH diplotypes across all chromosomes and mortality of COVID-19. Diplotypes with less than 10 cases were discarded. (b) UCSC genome browser view of the region containing the diplotype with a significant p-value in chromosome 4.

## Discussion

In this work, we introduced an efficient algorithm, ROH-DICE, for finding clusters of ROH regions in very large cohorts. The algorithm can find all clusters of ROH regions based on the given parameters: the minimum number of individuals, the minimum length of the ROH regions and the objective function. The running time of the algorithm is linear to the size of the genotype panel which enables fast processing of millions of individuals without requiring extravagant resources.

Using ROH-DICE, we conducted a systematic investigation of ROH diplotype clusters in a large population cohort, the UK Biobank. To the best of our knowledge, there has been no such investigation of the genomic distribution of LOH diplotypes conducted previously. We found over 1.8 million ROH diplotype clusters spanning over 100 SNPs and shared by over 100 individuals. While we reported this single data point, the interpretation of the genome-wide ROH diplotype distribution is difficult. First, the expected distribution of ROH diplotype clusters is not known. In general, such ROH regions may be under strong recessive selection, and thus the ROH cluster frequency might be higher under a null model (no selection). Further, the distribution of ROH diplotype clusters may depend on the demographic history. Therefore, further investigation with more data points and theoretical modeling is a research avenue wide open.

We found a strong association between non-cancerous diseases and some ROH diplotypes. The majority of ROH regions harboring strong associations with non-cancerous diseases were located in the extended HLA region in chromosome 6. As expected, most of the related diseases were also auto-immune system disorders. While the association signals we found mostly overlap with existing GWAS hits, we are testing different genetic effects. The existing GWAS are mainly testing the additive effects of single SNPs, while we are testing the recessive effects of relatively long haplotypes. In a sense, our analysis is similar to traditional family-based homozygosity mapping^12^, but at a population scale. Future works are warranted to fully develop this potential new gene mapping approach.

## Methods

### Identification of haplotype clusters in PBWT

Positional Burrows-Wheeler Transform (PBWT) proposed by Durbin^21^ facilitates an efficient approach to search for all pairs of long matches in haplotype or genotype panels. The basic idea behind the PBWT search is to sort the panel at each site by their reversed prefix order. As a result, the matches in the panel will be placed adjacent to each other. However, at the time we started this project, all existing PBWT algorithms^21,40–42^ were aimed at identifying pairwise matches. In this work, we propose to employ the PBWT data structures to search for clusters of multi-way matches instead of individual pairs of matches. Independent of our work, a couple of algorithms have been proposed to find haplotype blocks in a PBWT panel^43,44^. The algorithm by Cunha et al.^43^ may not be feasible to handle biobank scale data. The recent proposed algorithms by Alanko et al.^44^, however, will scale well for large-scale data, but they aim to enumerate all maximal haplotype blocks. For datasets comprising hundreds of thousands or millions of individuals, the number of reported clusters of any length may be excessive. Moreover, a minimum length threshold in terms of both sites and number of individuals would be more meaningful for downstream analysis especially association analysis, e.g. where a minimum number of cases is required. Hence, after detecting all possible clusters, filtering has to be applied to remove spurious clusters. Here, we formulate the haplotype blocks problem with two distinct objective functions which will reduce the complexity of filtering the detected clusters afterwards.

### Block maximal match problems

Based on the different formulations of the problem, we may have different objective functions: the first problem is to find all clusters with at least *L* sites that are shared among at least *t* sequences while maximizing the number of sequences for each cluster. Using proper data structures, we can keep track of the starting position of the matches and report them efficiently. The second problem is to find clusters with at least *L* sites among at least *t* sequences while maximizing the number of sites for each cluster. Again, the sequences that share a consensus are put in the same block.

PBWT sorting at the site *k* places sequences with identical reverse prefixes into clusters of matches which are adjacent to each other. We refer to these clusters as blocks, where the number of sequences *W* is the width of the block in terms of the number of haplotypes, and the length of matches *L* is the length of the block in terms of the number of sites. Recall the concept of set maximal match of Durbin^21^ as the pairwise haplotype match that cannot be extended at either end. We extend the concept of set maximal match to *block maximal match*, i.e., the haplotype match block that cannot be extended. As the block is a 2D object, the extension can be defined either lengthwise or widthwise. Therefore, we can define the *lengthwise block maximal match* as the matching block that cannot be extended lengthwise. Similarly, the *widthwise block maximal match* as that which cannot be extended widthwise.

For the PBWT block match problem, the goal is to identify all block maximal matches that have a minimal sequence length L and a minimal width W. Note that for an identified PBWT block, the boundary of the block may not be exactly defined (see **Figure 6** for an example). We can either report the block boundary that maximizes the length - *length-maximal PBWT block*, or the block boundary that maximizes the width - *width-maximal PBWT block*. We developed exact algorithms for identifying and reporting block maximal matches. This is achieved by using proper data structures tracking the starting position of the matches and the upper and lower boundaries of each matching block. A detailed description of the algorithms is provided in the cPBWT algorithms subsection.

**Figure 6:**
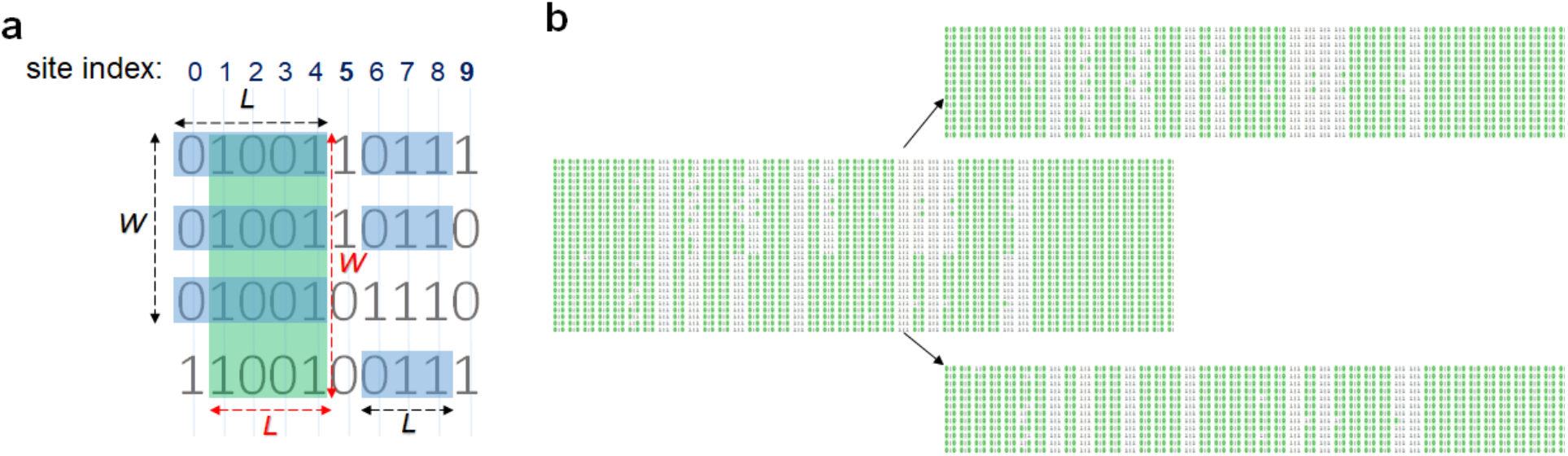
Consensuses of haplotype matches with a minimum length (*L*) of 3 and a minimum width (*W*) of 3. The green rectangle ending at site 4 highlights a cluster which meets the requirement of *W* >= 3 and *L* >= 3 while maximizing the number of individuals (width-maximal). The blue rectangles ending at 4 maximizes the number of sites (length-maximal). The blue rectangles ending at site 8 shows a cluster with *W* >= 3 and *L* >= 3 with maximizing the number of sites and/or number of individuals. Merging and splitting individuals using their ROH consensuses (b). Figure (b) shows a real example of two clusters (in chromosome 1 of UKBB) with the same starting and ending positions but different consensuses. Each line represents one individual and 0-alleles are colored in green. The individuals with the same ROH consensus while tolerating heterozygous sites are put in the same cluster.

### cPBWT algorithms

#### Maximizing the number of haplotypes

Given a haplotype or genotype panel, the objective is to find all matches greater than a given length *L* that are shared among at least *c* haplotypes (or individuals). By sorting the panel at each site the matches are placed in the same block. The divergence value for each sequence contains the starting position of the match to its preceding sequence in the reversed prefix order. The matches are separated by a sequence with the divergence value greater than *k - L*. In order to maximize the number of sequences, the maximum value of the divergence values in each block is considered. The size of the block should also be greater than *c* in order to be reported. Algorithm 1 (Supplementary Materials) illustrates the procedure for finding long matches while maximizing the number of haplotypes or sequences in detail. Algorithm 2 (Supplementary Materials) illustrates the procedure for updating the intermediate variables *V* and *Q* to compute *d*_*k*+*1*_ and *a*_*k*+*1*_ based on the *d*_*k*_ and *a*_*k*_.

#### Maximizing the length of match

The objective is to find the longest matches greater than a given length *L* shared among at least *c* sequences. Basically, the match will not be reported if the block of matches can be extended while at least *c* sequences are not terminating. To do this, two conditions should be held: First, at least *c* sequence for one allele should be present in the block, and second, the c-th lowest divergence value in the block should be greater or equal to the *c*-th lowest divergence of the matches ending with the allele with at least *c* occurrences. To find the *c*-th lowest divergence value, the Quickselect algorithm, a modified one-sided version of Quicksort^45^, is used. Quickselect has the average time complexity of *O*(*N*), where *N* denotes the size of the given list. Algorithm 3 (Supplementary Materials) illustrates the procedure of finding long matches while maximizing the length of match in detail.

### The ROH-DICE algorithm

Any of the two cPBWT algorithms can be applied to search for ROH-diplotype clusters from genotype data. ROH-DICE maps the genotype sequence *x*, defined over the alphabet of {0,1,2}, into a compressed haplotype sequence *y*, defined over the alphabet of {0,1}. For homozygous sites, the mapping is straightforward: for *x*_*i*_ = 0, *y*_*i*_ = 0; for *x*_*i*_ = 2, *y*_*i*_ = 1. For heterozygous sites *x*_*i*_ = 1, a random value from 0 and 1 was assigned with a probability of ½ for 0 and ½ for 1. The identified maximal matching blocks in the PBWT panel comprising all compressed haplotype sequences {*y*_*i*_}, correspond to the approximate ROH clusters in the original genotype sequences {*x*_*i*_}. The possible range for random assignment can be increased by the user. cPBWT algorithms are also capable of handling multi-allelic sites. Increasing the range of random assignment will decrease the tolerance of heterogeneity in the ROH regions. In this work, we simply considered bi-allelic random assignment.

### UK Biobank Data Set

The phased haplotype data of the UK Biobank (version 2) data comprising 658,720 sites were extracted. The Data-Field 20002 contains self-reported non-cancer illness comprising 445 categories (diseases). For the association analysis, 430,437 individuals with British ethnicity were selected. The ethnic backgrounds were extracted using the Data-Field 21000.

### Genetic association analysis

We computed the p-values for each disease in all detected ROH clusters that were present in at least 10 individuals. P-values were computed using chi-squared test considering the following numbers: D1: Number of individuals sharing a disease within the detected consensus of ROH. N1: Number of individuals in the detected ROH not sharing the disease. D2: Total number of individuals sharing the disease subtracting D1. N2: M – N1 – N2 – D2, where M denotes the total number of individuals. 100 regions with the lowest p-values (for any disease) were selected and further investigated using PHESANT (downloaded on August 22, 2018).

For PHESANT analysis, Age was calculated manually using the date of attending assessment centre (53), year of birth (34) and month of birth (52). Sex (31), Genetic principal components (22009), Number of self-reported non-cancer illnesses (135), Genotype measurement batch (22000) and Non-cancer illness (20002) fields were also maintained. All diplotypes in the 100 regions were considered as traits of interest. Most of the regions include multiple clusters with the same starting and ending positions but different consensus. We considered all of the clusters in the same region as traits of interest (660 traits of interests in total).

### Retrieval and annotation using the genetic association results from Neale Lab

Each of the associations (computed by PHESANT) was validated against the GWAS results published by Neale’s lab^b^. For each disease in each cluster (according to PHESANT), all reported SNPs within the genomic region of the cluster that were reported to be associated with the disease (according to Neale’s lab results) were searched and the SNP with the lowest p-value was reported.

### COVID-19 Mortality and ROH diplotypes

Two tables “covid19_result.txt” and “death.txt” provided by the UK Biobank were downloaded on July 24 2020. The table “covid19_result.txt” contains the test results whether the sample was reported as positive or negative for COVID-19. The table “death.txt” includes the date of death for samples. In the July 24, 2020 release of the table in UK Biobank, 201 British individuals have been reported COVID-19 test positive and died in 2020. Those individuals were considered as cases for mortality analysis. A total of 8120 British individuals have been tested for COVID-19. The controls contained the individuals who had been tested but no death information was provided for them. We tested all detected ROH diplotypes for COVID-19 mortality association (with at least 10 cases) using the Chi-square test. For the Chi-square test, the total number of individuals *M* corresponds to the number of tested individuals for COVID-19 (8120).

## Availability

The source code for our ROH-DICE methods is available at: https://github.com/ZhiGroup/roh-dice.

## Data Availability

Access to the UK Biobank Resource is available by application (https://www.ukbiobank.ac.uk).

## Acknowledgements

AN, SZ and DZ were supported by the National Institutes of Health grant R01 HG010086. AN and DZ were also supported by the National Institutes of Health grant OT2 OD002751. This research has been conducted using the UK Biobank Resource under Application Number 24247. We thank Dr. Irmgard Willcockson for proofreading.

## Footnotes

a http://www.nealelab.is/blog/2017/9/15/heritability-of-2000-traits-and-disorders-in-the-uk-biobank

b http://www.nealelab.is/blog/2017/9/15/heritability-of-2000-traits-and-disorders-in-the-uk-biobank, Accessd Jul 27, 2018

