## Supplementary Materials for "Discovery of runs-of-homozygosity diplotype clusters and their associations with diseases in UK Biobank"

**Algorithm 1** Find all blocks of matches while maximizing the number of haplotypes

```
procedure MAXIMAIZE_HAPLOTYPES(x_k, t, a_k, d_k, L, c)
    V = [] , Q = []
    for r = 0 to t do
        m.append([])
        pq.append(k+1)
        V.append([])
        Q.append([])
    end for
    i0 ← 0
    for i = 0 to M − 1 do
        if d_k[i] > k − L + 1 then
            changed ← false
            for p = 0 to t do
                for q = p + 1 to t do
                    if len(m[p]) > 0 and len(m[q] > 0) then
                        changed ← true
                        break
                    end if
                end for
            end for
            if changed and i - i0 >= c then
                blockMatches = []
                d_min ← 0
                blockMatches.append[a_k[i0]]
                for i_b = i0 + 1 to i do
                    blockMatches.append([ak[i_b]])
                    if (d_k[i_b] > d_min) then
                        d_min = div[i_b]
                    end if
                end for
                report d_min and blockM atches
            end if
            i0 ← i , m = []
            for r = 0 to t do
                m.append([])
            end for
        end if
        updateV_Q(x_k[l], i, t, a_k, d_k, pq, V, Q)
    end for
    a_k+1 ← concatenate[V0, V1, ..Vt]
    d_k+1 concatenate[Q0, Q1, ..Qt]
end procedure
```

**Algorithm 2** Update intermediate variables for cPBWT

```
procedure UPDATEV_Q(allele, i, t, a_k, d_k, pq, V, Q)
    for p = 0 to t do
        if d_k[i] > pq[p] then
            pq[p] ← d_k[i]
        end if
    end for
    V [allele].append(a_k[i])
    for p = 0 to t do
        if allele == p then
            Q[allele].append(pq[allele])
            pq[allele] ← 0
            mab[p].append(a_k[i])
        end if
    end for
end procedure
```

**Algorithm 3** Find all blocks of matches larger than $L$ while maximizing the length of match

```
procedure MAXIMAIZE HAPLOTYPES(x_k, t, a_k, d_k, L, c)
    V = [] , Q = []
    for r = 0 to t do
        m.append([])
        pq.append(k+1)
        V.append([])
        Q.append([])
    end for
    i_0 ← 0
    for i = 0 to M − 1 do
        if d_k[i] > k − L + 1 then
            changed ← false
            for p = 0 to t do
                for q = p + 1 to t do
                    if len(m[p]) > 0 and len(m[q] > 0) then
                        changed ← true
                        break
                    end if
                end for
            end for
            if changed and i - i_0 >= c then
                blockMatches = [], d_min ← 0, f_a ← a_k[i_0], f_d ← d_k[i_0 + 1]
                T_d[f_a].append(f_d), all_d.append(f_d)
                for i_a = i_0 + 1 to i do
                    blockMatches.append([a_k[i_b]])
                    if (d[i_a] > d_min) then
                        d_min = d_k[i_b]
                    end if
                    f_a ← x_k[a_k[i_a]], T_d[f_a].append(d_min), all_d.append(d_min)
                end for
                for r = 0 to t do
                    if len[T_d[r]] >= c and quick_select(T_d[r], c − 1) <= quick_select(all_d, c − 1) then
                        skip

                        i_a ← i_0
                        if f_d <= quick select(all_d, c − 1)) then
                            blockMatches.append([a_k[i_0]]
                            for i_a = i_0 + 1 to i do
                                if d_k[i_a] <= quick_select(all_d, c − 1) then
                                    blockMatches.append(a_k[i])
                                end if
                            end for
                            d_r = quick select(all_d, c − 1)
                            report blockMatches and d_r

                        end if
                        i_0 ← i , m = []
                        for r = 0 to t do
                            m.append([])
                        end for
                    end if
                    updateVQ(x_k[l], i, t, a_k, d_k, pq, V, Q)
                end for
                a_{k+1} ← concatenate[V_0, V_1, ..V_t]
                d_{k+1} ← concatenate[Q_0, Q_1, ..Q_t]
```
